# Longitudinal interaction between muscle impairments and gait pathology in growing children with Duchenne muscular dystrophy

**DOI:** 10.1101/2024.11.27.24318103

**Authors:** Ines Vandekerckhove, Geert Molenberghs, Marleen Van den Hauwe, Nathalie Goemans, Liesbeth De Waele, Anja Van Campenhout, Friedl De Groote, Kaat Desloovere

## Abstract

**Aim:** The aim of this longitudinal observational cohort study was to investigate the longitudinal interaction between progressive muscle impairments and progressive gait pathology in growing boys with Duchenne muscular dystrophy (DMD).

**Methods:** Thirty-one boys with DMD (aged 4y7mo-16y5mo), were repeatedly measured between 2015 and 2022, resulting in a total dataset of 152 observations. Fixed dynamometry, goniometry and 3D gait analysis were used to asses lower limb muscle weakness, passive range of motion and gait. Joint random-effects models between gait and muscle impairment outcomes were fitted. The correlation between the random intercepts (*r_a_*) and random slopes (*r_b_*) indicated the degree to which respectively the starting levels and progression rates over time of two outcomes were related in boys with DMD.

**Results:** Specific muscle impairments were related to specific gait features, in terms of starting levels (*r_a_*=0.470-0.757; p<0.029) and progression rates (*r_b_*=0.547-0.812; p<0.024).

**Interpretation:** The findings improved insights into how specific muscle impairments may contribute to specific gait features in DMD. This could enhance clinical decision making, advance rehabilitation, orthotic and orthopedic interventions, and reveal sensitive outcome measures to prove the efficacy of novel treatments in clinical trials.

**What this paper adds:**

- Objective quantification of the relationships between muscle impairments and gait features
- Relationships between starting levels of specific muscle impairments and specific gait features
- Relationships between progression rates of specific muscle impairments and specific gait features
- Improved insights into how muscle impairments may contribute to gait features

## Introduction

Duchenne muscular dystrophy is a severe X-linked neuromuscular disorder, affecting one in 3,500-6,000 newborn boys^1–3^. Disrupted dystrophin function due to mutations in the dystrophin gene causes progressive muscle degeneration with loss of contractile tissue and replacement by fat and fibrotic tissue^2^. Consequently, boys with DMD experience progressive muscle weakness and contractures that contribute to altered posture and gait, with eventually loss of ambulation between 7.1 and 18.6 years of age (mean age: 12.7 years)^1,2,4^. Delaying loss of ambulation is one of the treatment goals in DMD, in order to maintain a certain level of functionality and to postpone spinal deformities and contractures^2^. However, orthopedic and orthotic treatments that aim at optimizing gait performance indicated conflicting results on gait decline^5–9^ and clinical development of promising novel treatments has been hindered due to lack of sensitive outcome measures^10,11^. An improved understanding of how the progress in underlying muscle impairments is associated with increasing gait pathology in DMD can improve clinical decision making, lead to progress in rehabilitation, orthotic and orthopedic interventions, and reveal sensitive outcome measures to prove the efficacy of novel treatments in clinical trials.

Despite the clinical consensus that muscle impairments alter gait in DMD, insight in specific associations between muscle impairments and gait characteristics is lacking. The relationship between underlying muscle impairments and clinical assessments, such as the 6-min walk test, the North Star Ambulatory Assessment, and timed test has been extensively studied. However, these clinical assessments only measure global gross motor function and are characterized by high variability in test outcomes^12^. Investigating the association with the gait pattern measured in detail by 3D gait analysis (3DGA) instead could reveal new and important insights. For example, it has been hypothesized that tiptoeing gait is a compensation mechanism for knee extension weakness, as it positions the ground reaction force close to or in front of the knee joint center resulting in smaller knee extension moments^1,7,13–15^. To date, only one study has investigated the relationship between muscle weakness and the gait pattern in DMD, but no meaningful associations were found^16^. However, the latter study was based on cross-sectional data of 15 boys with DMD. Due to the progressive and heterogeneous nature of DMD, these associations should be estimated from larger sample sizes and longitudinal data.

Linking the progression in muscle impairments to the progression in gait pathology in growing children with DMD may highlight the potential role of impairments on gait deterioration. We recently established longitudinal trajectories of muscle impairments^17^ and gait features^18^ through an extensive 5-7 year follow-up. The muscle impairments were expressed as deficits in reference to TD peers, allowing the established trajectories to reflect pathological changes. The pathological trajectories of most muscle impairments showed a non-linear, piecewise pattern, characterized by an initial phase of stability lasting until 6.6-9.5 years, and a subsequent decline after these ages. Muscle weakness of all measured lower-limb muscles and ankle dorsiflexion range of motion exhibited the steepest declines, resulting in large deficits at older ages. The progressive gait pathology evolved towards more anterior pelvic tilt, hip flexion, internal foot progression and less dorsiflexion at initial contact. There was a high inter-subject variability in the longitudinal trajectories of both muscle impairments and gait features, but it remains unclear which muscle impairments are related to which gait features and how their progression rates relate.

The aim of this study was to investigate the longitudinal interaction between progressive muscle impairments and progressive gait features in growing boys with DMD. Specifically, we aimed to determine whether the starting levels of specific muscle impairments is related to the starting levels of specific gait features and whether their progression rates are related. This is necessary to improve insights into how specific muscle impairments contribute to specific gait features.

## Methods

### Participants

We conducted a longitudinal observational cohort study between 2015 and 2022. The study protocol consisted of multivariate repeated assessments with a varying number of assessments among participants and varying time intervals between assessments.

Boys with DMD were recruited via the Neuromuscular Reference Centre (NMRC) in the University Hospital Leuven. The study included boys with a confirmed genetic diagnosis of DMD, who were between 3 and 16 years old at baseline, and able to walk at least 100 meters. Exclusion criteria encompassed a clinical presentation of Becker muscular dystrophy, any history of muscle lengthening surgery, and cognitive or behavioral disorders that impeded accurate measurements. Corticosteroids intake and clinical trial participation with disease-modifying medication were allowed. NMRC implements a proactive and preventive approach to manage contractures with an early introduction to night-time ankle foot orthosis (AFOs), often alongside the start of corticosteroids, before contracture development, and occasional serial casting if early losses in ankle dorsiflexion range of motion still appear.

This study was approved under the Declaration of Helsinki by the local ethics committee (Ethical Committee UZ Leuven/KU Leuven; S61324). All methodology adhered to the relevant regulations and guidelines. Written informed consents were obtained from the parents or participants’ caregivers, and participants aged 12 years or older provided informed assents.

### Data collection and analysis

Anthropometric measures, i.e., body mass, height and lower limb segment lengths, were collected at each observation. Muscle weakness was measured unilaterally on the weakest side, which was determined based on the manual muscle testing. Contractures and gait deviations were measured bilaterally. However, only the measures on the weakest side were further included in the analyses to be consistent with the muscle weakness measurements. The assessed side was randomly selected, if no weakest side could be determined.

We made a selection of muscle impairments and gait features based on our previous longitudinal analysis (Table 1)^17–19^. Gait features that showed the most interesting trajectories were included. The selection of muscle impairments for the analyses was defined through clinical reasoning, where we focused on muscle impairments that were previously postulated as potential explanations for the notable gait features.

**Table 1:** Overview of the selected gait outcomes, the reasoning behind their selection, and the impairments that are believed to be related to these gait outcomes.

| Selected gait parameters | Reason | Impairments postulated to relate to gait parameters |
| --- | --- | --- |
| Minimal anterior trunk angle (°) | Clinically relevant decrease in more severely affected gait patterns compared to mildly affected gait pattern<sup>19</sup> | Hip extension weakness |
| Lateral trunk range of motion (°) | Clinically relevant increase in more severely affected gait patterns compared to mildly affected gait pattern<sup>19</sup> | Hip abduction strength deficit |
| Maximal anterior pelvic tilt angle (°) | Previously reported significant longitudinal increase<sup>30</sup> | Hip extension strength deficit<br>Knee extension strength deficit |
| Dorsiflexion angle at initial contact (°) | Previously reported significant longitudinal decrease<sup>30</sup> | Knee extension strength deficit<br>Dorsiflexion strength deficit<br>Dorsiflexion range of motion deficit |
| Maximal dorsiflexion angle during swing (°) | Previously reported significant interaction effect between time and baseline age<sup>30</sup> | Dorsiflexion strength deficit<br>Dorsiflexion range of motion deficit |
| Maximal internal foot progression angle during stance (°) | Previously reported significant longitudinal increase<sup>30</sup> | Poorly understood |
| Maximal hip extension moment during stance (Nm/kg) | Previously reported significant longitudinal decrease<sup>30</sup> | Hip extension strength deficit |
| Maximal knee extension moment during stance (Nm/kg) | Clinically relevant decrease in more severely affected gait patterns compared to mildly affected gait pattern<sup>19</sup> | Knee extension strength deficit |
| Minimal knee power during burst K1 (W/kg) | Clinically relevant decrease in more severely affected gait patterns compared to mildly affected gait pattern<sup>19</sup> | Knee extension strength deficit |
| Normalized step width (/) | Previously reported significant longitudinal increase as well as interaction effect between time and baseline age<sup>30</sup> | Hip abduction strength deficit |

kg, kilogram; Nm, Newton meter; W, Watt;

#### Muscle weakness

Hip extension, hip abduction, knee extension, and ankle dorsiflexion muscle weakness were assessed with an instrumented strength assessment^20,21^. Hereto, maximal voluntary isometric contractions (MVIC) were performed using a fixed dynamometer (MicroFet, Hogan Health Industries, West Jordan, UT United States) in a standardized test position. The mean maximal force over one to three representative MVIC trials was multiplied with its lever arm with respect to the joint to calculate the mean maximal joint torque per muscle group. Mean maximal joint torques were converted into unit-less z-scores using anthropometric-related TD percentile curves for muscle strength (n=153)^22^. As this accounts for typical strength development, these z-scores reflect muscle strength deficits with respect to TD peers.

#### Contractures

We included only the plantar flexion contractures in further analysis because our previous work^17^ highlighted that this was the only contracture with a clear longitudinal trajectory. The passive range of motion (ROM) of ankle dorsiflexion with knee extended and knee flexed in 90°^23^ was measured with goniometry in degrees during a standardized clinical examination. Passive ROM measures were converted into unit-less z-scores using the age-related normative reference values of Mudge et al.^23^, as previously described^17^. As this accounts for typical reduction in passive ROM, these z-scores reflect ROM deficits with respect to TD peers.

#### Gait

Gait was measured by 3D gait analysis according to a previously described protocol^18,19^. The boys with DMD walked barefoot at self-selected speed on a 10-meter walkway. The Plug-In Gait Full-body reflective marker (diameter: 14 mm) model was applied. Marker trajectories were recorded with a 10-15 Vicon camera system (Vicon-UK, Oxford, UK; sampling frequency of 100 Hz; built-in Woltring filter with mode MSE and smoothing of 15 mm^2^) and ground reaction forces were captured with two embedded force plates (AMTI, Watertown, MA, USA; sampling frequency: 1500 Hz). Kinetic data was obtained by combining marker trajectories with ground reaction forces. Initial contacts and toe offs were manually indicated, using force plate data when available, to define gait cycles (GCs) in the Nexus software (Nexus 2.10. Vicon-UK, Oxford, UK). Subsequently, trunk, pelvis and lower limb kinematic waveforms (expressed in degrees), and waveforms for lower limb internal joint moments (expressed in Newton meters per kilogram body mass), and power (expressed in Watt per kilogram body mass) were estimated. Ten GCs with kinematic data, of which three to five GCs with kinetic data, were collected bilaterally. We examined the quality of collected GCs in a custom-made software in MATLAB (The Mathworks Inc., Natick, M.A., 2016 and 2021b). Kinematic and kinetic waveforms were resampled to 101 data points per GC. Spatiotemporal parameters were normalized as previously described^18,19,24^. Normalized spatiotemporal parameters, and the kinematic and kinetic waveforms of the selected GCs with good quality were averaged per observation separately for the right and left sides. Predefined gait features were obtained by calculating minima, maxima, range of motions, and values at specific events in the GC from the average continuous kinematic and kinetic waveforms. Only the gait features on the weakest side were further included.

### Statistical analysis

Joint random-effect models were applied to investigate the longitudinal correlation among multivariate outcomes. The outcome parameters were selected based on our previous results^17–19^ and pairs of muscle impairments and gait features to jointly model were chosen based on clinical reasoning (Table 1). A mixed model was first fitted on each individual outcome, according to the previously documented workflow^18,25^. Time course of the observations, starting from the baseline observation and calculated by subtracting baseline age from the age at each observation, was selected as the fixed effect. A random intercept modelled the variability in starting point, while a random slope for time modelled the variability in trajectory among subjects. Then, joint models with observation *j* in subject *i* to estimate gait_ij_ outcome and muscle_impairments_ij_ outcome were created by linking the random effects of the two outcomes as follows^26^:

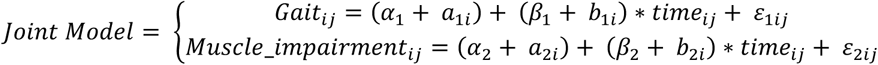

with α_1_=intercept of gait outcome; a_1i_=random intercept of gait outcome; β_1_=regression slope for time of gait outcome; b_1i_=random slope of β_1_; ε_1ij_=measurement error of gait outcome; α_2_=intercept of muscle impairment outcome; a_2i_=random intercept of muscle impairment outcome; β_2_=regression slope for time of muscle impairment outcome; b_2i_=random slope of β_2._ The random-effects (a_1i_, a_2i_, b_1i_,b_2i_) are assumed to be zero-mean normally distributed with variance-covariance matrix:

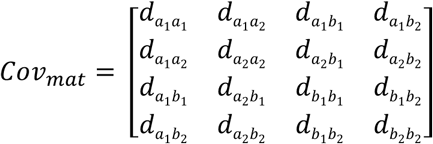

Further, the measurement error terms ε_1ij_ and ε_2ij,_ for gait and muscle impairment, respectively, are assumed independent, and independent from the random effects, with zero mean.

The estimates of the individual mixed models were used as starting values for the joint models, while unknown estimates were initiated at zero. Pearson correlations between random intercepts (*r_a_*) on the one hand and between random slopes (*r_b_*) on the other hand, were estimated using the expressions^26^:

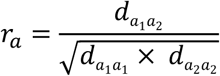

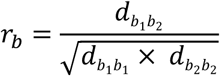

*r_a_* and *r_b_* indicate the degree to which respectively the starting levels and progression rates over time of two outcomes are related. Because correlations are bounded, testing for significance and calculation of 95% confidence intervals are conveniently estimated at Fisher z’s transformed scale. Upon calculation, they can be back transformed to the correlation scale. Correlation coefficients were interpreted as negligible (*r*<0.300), low (*r*=0.300-0.499), moderate (*r*=0.500-0.699), high (*r*=0.700-0.899), or very high (*r*≥0.900)^27^. All analyses and visualizations were conducted in SAS® (version 9.4, Statistical Analysis Software, SAS Institute Inc., Cary, NC, USA).

## Results

Thirty-one boys with DMD, aged between 4y7mo and 16y5mo, were repeatedly measured between 2015 and 2022 at multiple time points (median=3; Q1-Q3=2-5.5; min-max=1-10) with a median time interval of 0.51 years (Q1-Q3=0.49-1.01 years; min-max=0.4-3.5 years), covering a median follow-up time of 2.2 years (Q1-Q3=0.5-3.5 years; max=6.6 years). In total, the included dataset consisted of 152 measurement sessions (Appendix S1). All patient characteristics are documented in Table 2. The flowchart in Appendix S2 provides an overview of the collected data, the excluded data after quality check, the missing data, and the included data.

**Table 2:**
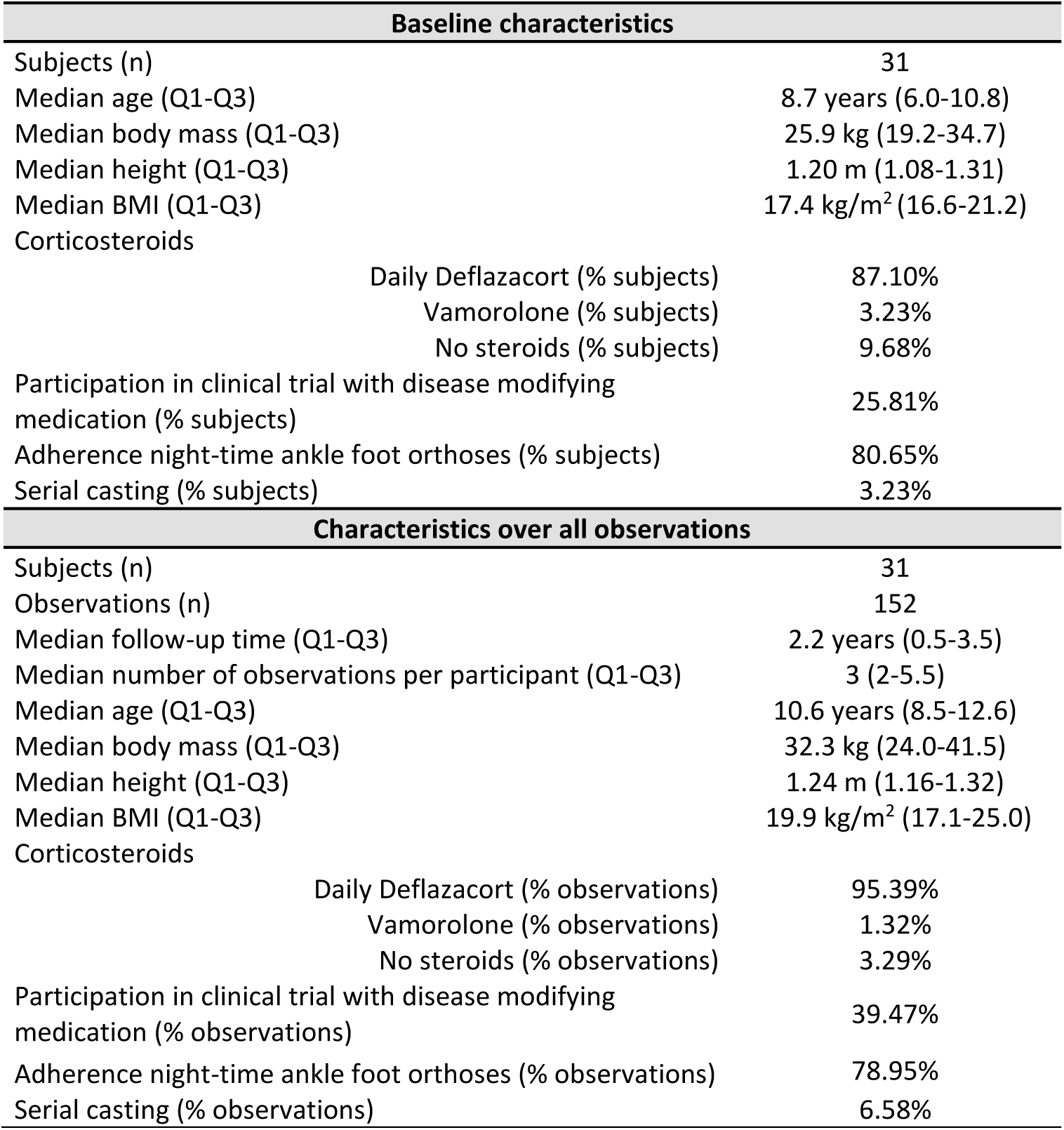
Group demographics.

The rate of decrease in minimal anterior trunk angle was moderately associated with the rate of decrease in hip extension strength (*r_b_*=0.586; p=0.0004; Table 3; Figure 1-a). Increased maximal anterior pelvic tilt angle was moderately associated to reduced hip extension strength (*r_a_*=-0.543; p=0.0040; Table 3; Figure 1-c), while the rate of increase in maximal anterior pelvic tilt angle was highly correlated with the rate of decrease in hip extension strength (*r_b_*=-0.812; p<0.0001; Table 3; Figure 1-c). Increased maximal anterior pelvic tilt angle was only minimally associated to reduced knee extension strength (*r_a_*=-0.470; p=0.0036; Table 3; Figure 1-d). The rate of decrease in dorsiflexion angle at initial contract was moderately associated with the rate of decrease in knee extension strength (*r_b_*=0.684; p=0.0237; Table 3; Figure 2-a) and in dorsiflexion range of motion with knee extended (*r_b_*=0.627; p=0.0202; Table 3; Figure 2-c). Decreased dorsiflexion angle at initial contact was only minimally associated with reduced dorsiflexion strength (*r_a_*=0.494; p=0.0113; Table 3; Figure 2-b), while it was highly associated with diminished dorsiflexion range of motion with knee extended (*r_a_*=0.732; p<0.0001; Table 3; Figure 2-c). Decreased maximal dorsiflexion angle during swing was minimally and moderately associated with reduced dorsiflexion strength (*r_a_*=0.494; p=0.0024; Table 3; Figure 3-a) and decreased dorsiflexion range of motion with knee extended (*r_a_*=0.663; p<0.0001; Table 3; Figure 3-b), respectively. The rate of increase in maximal internal foot progression angle was moderately associated with the rate of decrease in hip abduction strength (*r_b_*=-0.547; p=0.0117; Table 3; Figure 3-c). Reduced maximal hip extension moment was moderately associated with decreased hip extension strength (*r_a_*=0.536; p=0.0289; Table 3; Figure 4-a). Reduced maximal knee extension moment (*r_a_*=0.702; p<0.0001; Table 3; Figure 4-b) and increased minimal knee power during the first burst (*r_a_*=-0.757; p<0.0001; Table 3; Figure 4-c) were highly associated with diminished knee extension strength. Increased normalized step width was moderately associated with decreased hip abduction strength (*r_a_*=-0.547; p=0.0040; Table 3; Figure 4-d).

**Figure 1:**
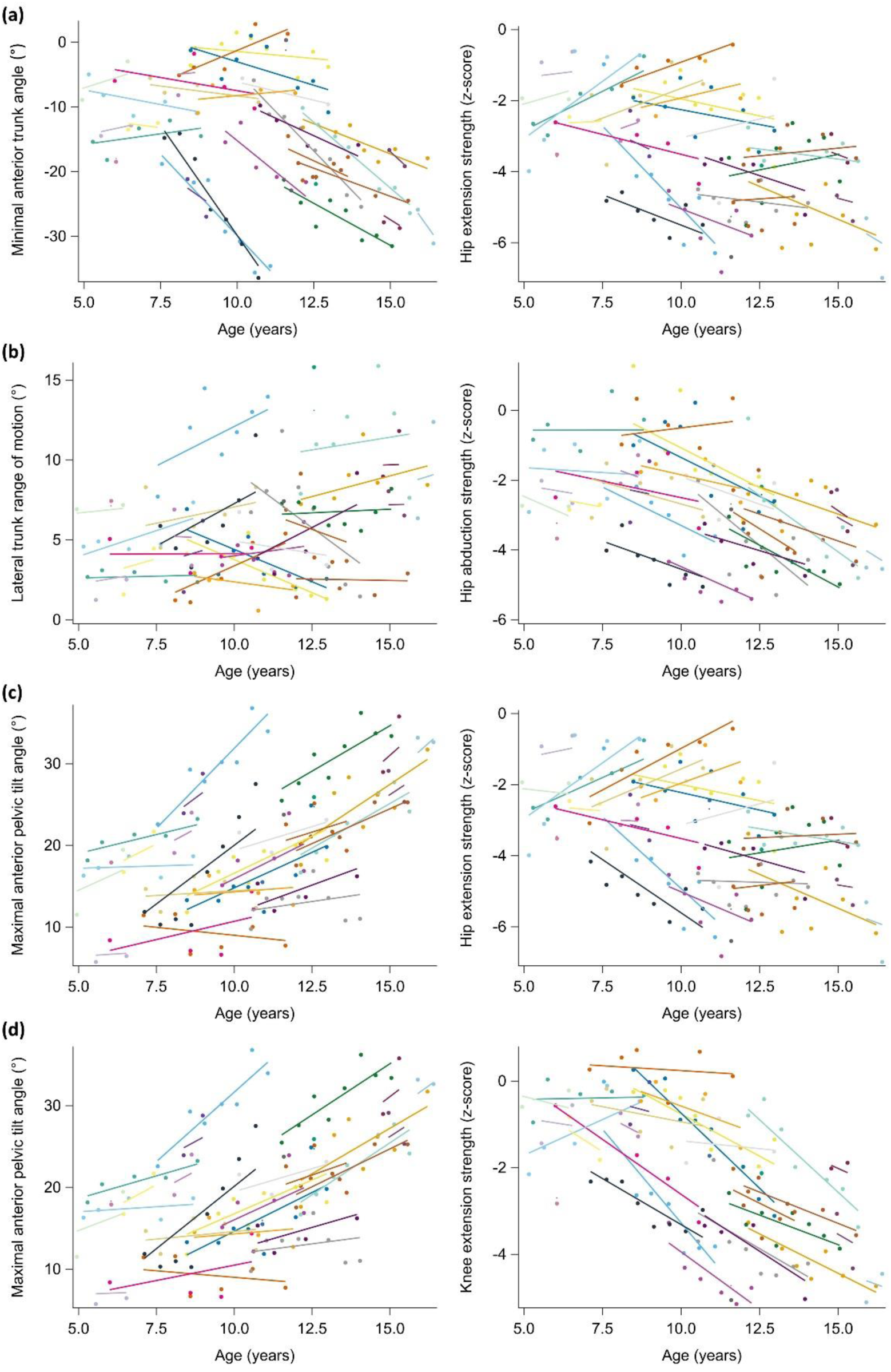
Relationship of starting levels and progression rates between two outcomes: minimal anterior trunk angle and hip extension strength **(a)**, lateral trunk range of motion and hip abduction strength **(b)**, maximal anterior pelvic tilt angle and hip extension strength **(c)**, and maximal anterior pelvic tilt angle and knee extension strength **(d).** Individual predicted profiles (colored lines) and actual observed outcomes (colored dots) are displayed. Each subject with DMD is indicated in a different color. Each panel illustrates the degree to which the starting points of the individual predicted profiles (i.e., correlation random intercepts) and the progression rates of the individual predicted profiles (correlation random slopes) are related between the two outcomes across subjects with DMD. The correlation coefficients are given in Table 3. Appendices S3-S4 provide the estimates for the fixed effects and the random-effects covariance matrix. DMD, Duchenne muscular dystrophy

**Figure 2:**
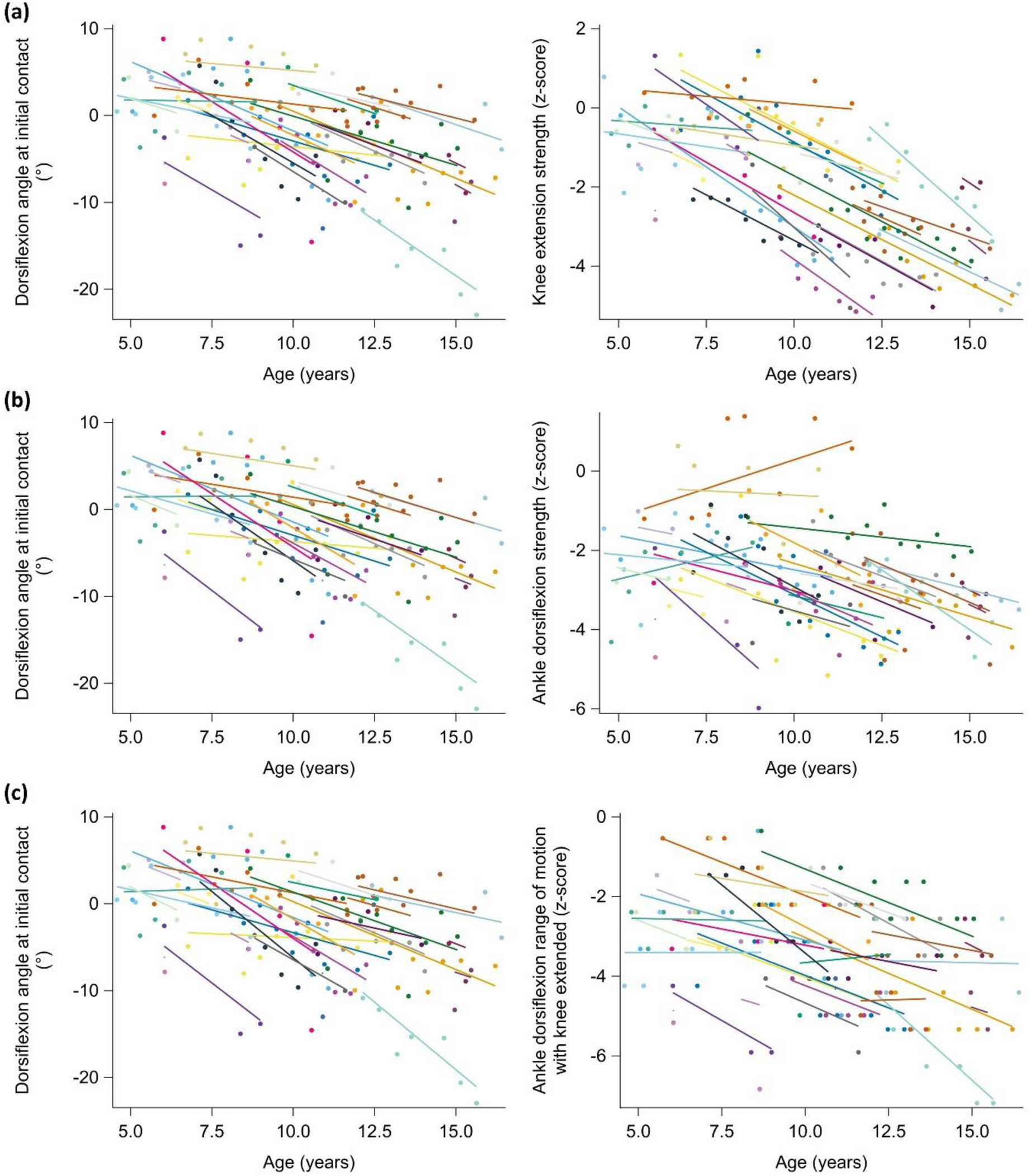
Relationship of starting levels and progression rates between two outcomes: dorsiflexion angle at initial contact and knee extension strength **(a)**, dorsiflexion angle at initial contact and ankle dorsiflexion strength **(b)**, and dorsiflexion angle at initial contact and ankle dorsiflexion range of motion with knee extended **(c)**. Individual predicted profiles (colored lines) and actual observed outcomes (colored dots) are displayed. Each subject with DMD is indicated in a different color. Each panel illustrates the degree to which the starting points of the individual predicted profiles (i.e., correlation random intercepts) and the progression rates of the individual predicted profiles (correlation random slopes) are related between the two outcomes across subjects with DMD. The correlation coefficients are given in Table 3. Appendices S3-S4 provide the estimates for the fixed effects and the random-effects covariance matrix. DMD, Duchenne muscular dystrophy

**Figure 3:**
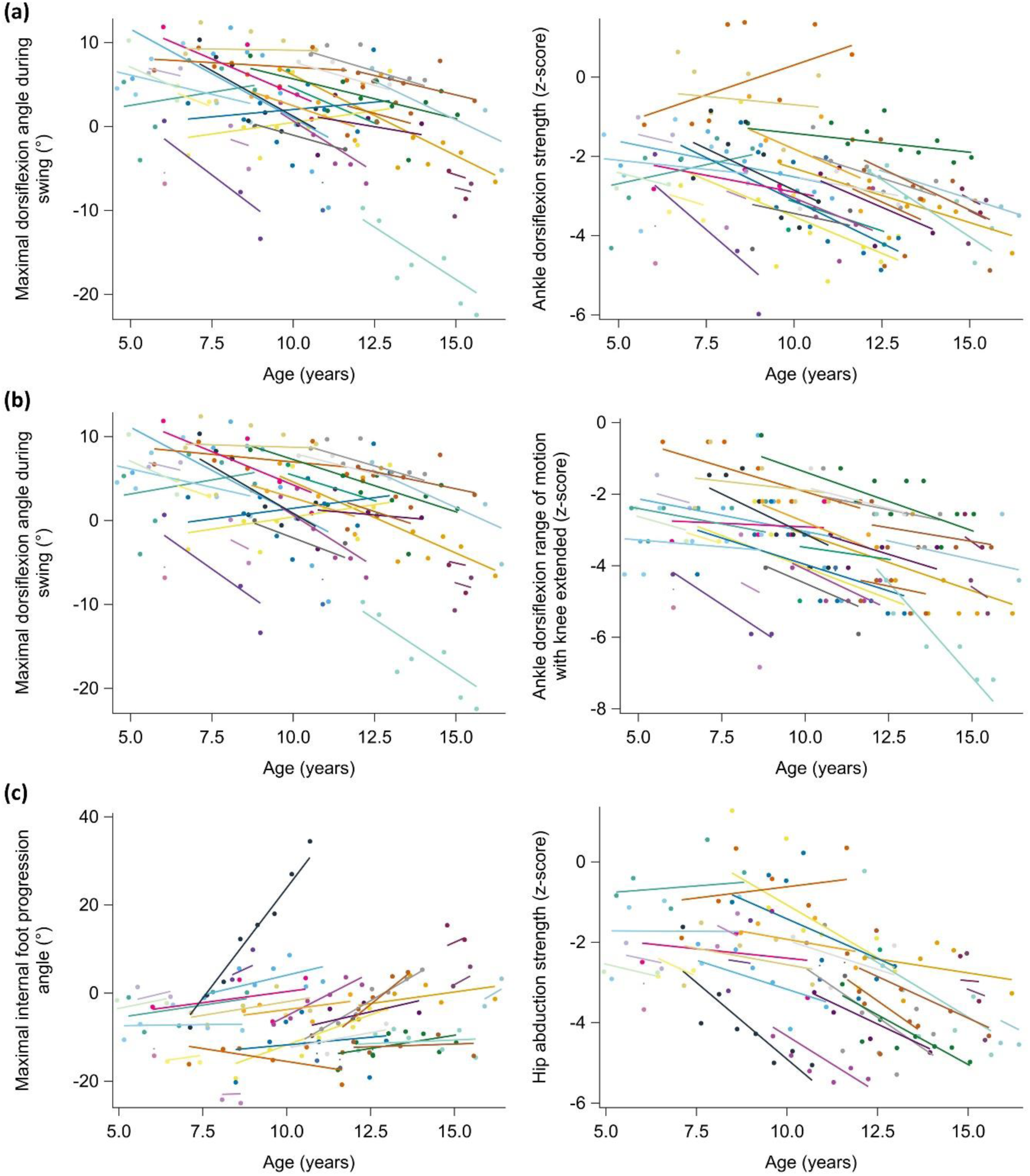
Relationship of starting levels and progression rates between two outcomes: maximal dorsiflexion angle during swing and ankle dorsiflexion strength deficit **(a)**, maximal dorsiflexion angle during swing and ankle dorsiflexion range of motion with knee extended **(b)**, and maximal internal foot progression angle during stance and hip abduction strength **(c)**. Individual predicted profiles (colored lines) and actual observed outcomes (colored dots) are displayed. Each subject with DMD is indicated in a different color. Each panel illustrates the degree to which the starting points of the individual predicted profiles (i.e., correlation random intercepts) and the progression rates of the individual predicted profiles (correlation random slopes) are related between the two outcomes across subjects with DMD. The correlation coefficients are given in Table 3. Appendices S3-S4 provide the estimates for the fixed effects and the random-effects covariance matrix. DMD, Duchenne muscular dystrophy

**Figure 4:**
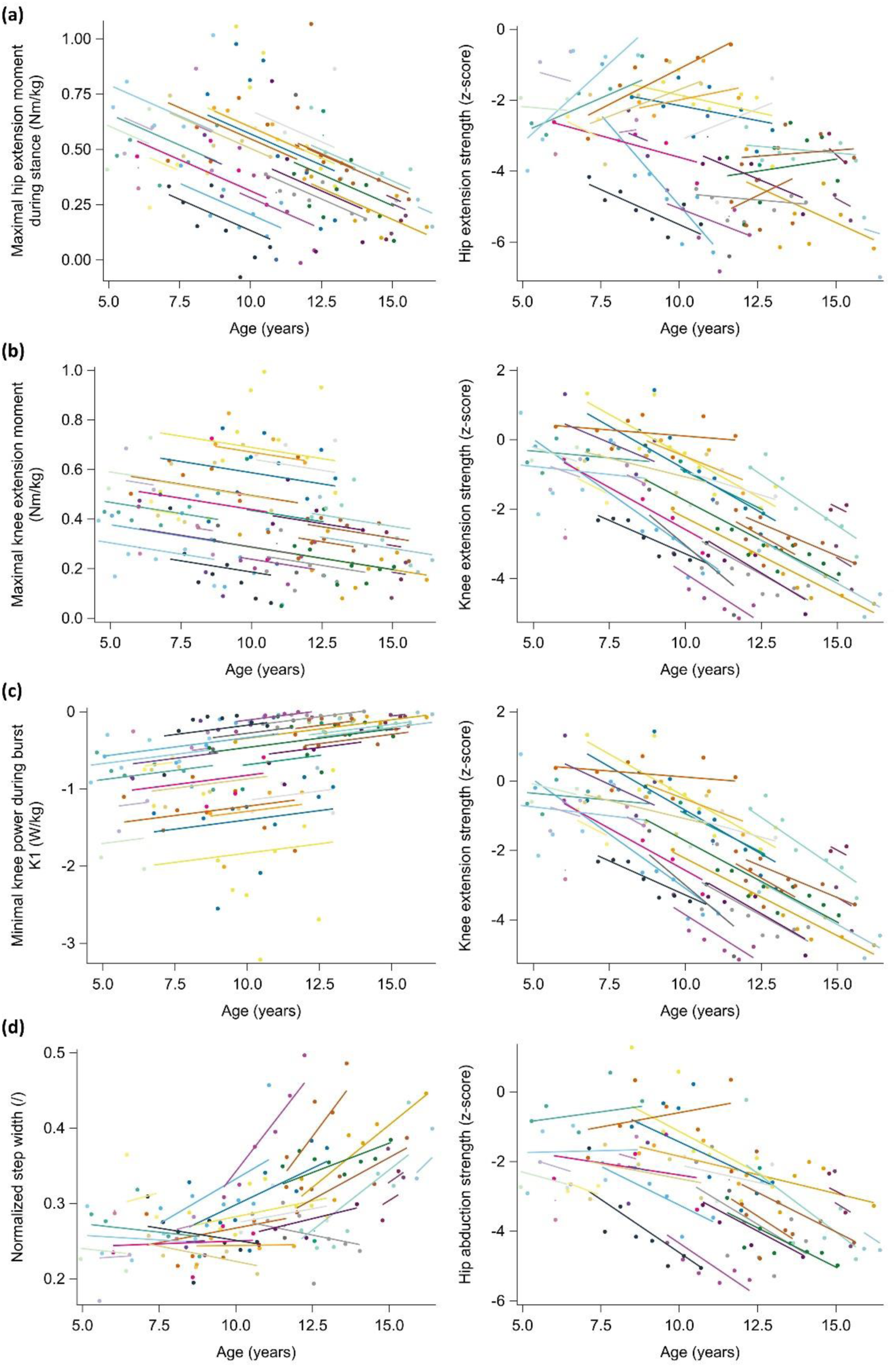
Relationship of starting levels and progression rates between two outcomes: maximal hip extension moment during stance and hip extension strength **(a)**, maximal knee extension moment during stance and knee extension strength **(b)**, minimal knee power during burst K1 and knee extension strength **(c)**, and normalized step width and hip abduction strength **(d)**. Individual predicted profiles (colored lines) and actual observed outcomes (colored dots) are displayed. Each subject with DMD is indicated in a different color. Each panel illustrates the degree to which the starting points of the individual predicted profiles (i.e., correlation random intercepts) and the progression rates of the individual predicted profiles (correlation random slopes) are related between the two outcomes across subjects with DMD. The correlation coefficients are given in Table 3. Appendices S3-S4 provide the estimates for the fixed effects and the random-effects covariance matrix. DMD, Duchenne muscular dystrophy; kg, kilogram; Nm, Newton meter; W, Watt;

**Table 3:** Correlation of random intercepts and random slopes of joint models between gait features and impairments in boys with DMD.

| Gait | Impairments | Correlation random intercepts | Correlation random slopes |
| --- | --- | --- | --- |
| | | $r_a$ [CI]<br>$p$-value | $r_b$ [CI]<br>$p$-value |
| Minimal anterior trunk angle (°) | Hip extension strength deficit (z-score) | 0.377 [-0.109; 0.717]<br>**0.1247** | 0.586 [0.292; 0.779]<br>**0.0004\*** |
| Lateral trunk range of motion (°) | Hip abduction strength deficit (z-score) | -0.187 [-0.484; 0.149]<br>**0.2752** | 0.529 [-0.111; 0.859]<br>**0.0995** |
| Maximal anterior pelvic tilt angle (°) | Hip extension strength deficit (z-score) | -0.543 [-0.771; -0.192]<br>**0.0040\*** | -0.812 [-0.922; -0.579]<br>**<0.0001\*** |
| Maximal anterior pelvic tilt angle (°) | Knee extension strength deficit (z-score) | -0.470 [-0.693; -0.165]<br>**0.0036\*** | -0.524 [-0.835; 0.039]<br>**0.0664** |
| Dorsiflexion angle at initial contact (°) | Knee extension strength deficit (z-score) | 0.200 [-0.1875; 0.534]<br>**0.3111** | 0.684 [0.111; 0.916]<br>**0.0237\*** |
| Dorsiflexion angle at initial contact (°) | Dorsiflexion strength deficit (z-score) | 0.494 [0.122; 0.744]<br>**0.0113\*** | 0.568 [-0.038; 0.868]<br>**0.0643** |
| Dorsiflexion angle at initial contact (°) | Dorsiflexion range of motion deficit (z-score) | 0.732 [0.564; 0.841]<br>**<0.0001\*** | 0.627 [0.114; 0.876]<br>**0.0202\*** |
| Maximal dorsiflexion angle during swing (°) | Dorsiflexion strength deficit (z-score) | 0.494 [0.189; 0.712]<br>**0.0024\*** | 0.385 [-0.239; 0.784]<br>**0.2210** |
| Maximal dorsiflexion angle during swing (°) | Dorsiflexion range of motion deficit (z-score) | 0.663 [0.402; 0.825]<br>**<0.0001\*** | 0.409 [-0.185; 0.784]<br>**0.1705** |
| Maximal internal foot progression angle during stance (°) | Hip abduction strength deficit (z-score) | -0.291 [-0.539; 0.003]<br>**0.0527** | -0.547 [-0.798; -0.136]<br>**0.0117\*** |
| Maximal hip extension moment during stance (Nm/kg) | Hip extension strength deficit (z-score) | 0.536 [0.061; 0.813]<br>**0.0289\*** |  |
| Maximal knee extension moment during stance (Nm/kg) | Knee extension strength deficit (z-score) | 0.702 [0.482; 0.838]<br>**<0.0001\*** |  |
| Minimal knee power during burst K1 (W/kg) | Knee extension strength deficit (z-score) | -0.757 [-0.870; -0.566]<br>**<0.0001\*** |  |
| Normalized step width (/) | Hip abduction strength deficit (z-score) | -0.547 [-0.775; -0.194]<br>**0.0040\*** | -0.397 [-0.763; 0.161]<br>**0.1572** |

The correlation of random intercepts indicates the degree to which the starting points of the individual predicted profiles are related between two outcomes across subjects with DMD. The correlation of random slopes indicates the degree to which the progression rates of the individual predicted profiles are related between two outcomes across subjects with DMD. $p$ -values in bold indicate significance level at $p < 0.05$ . The colors indicate the strength of the relationship: dark green, high; middle green, moderate; light green, low. CI, 95% confidence interval; DMD, Duchenne muscular dystrophy; kg, kilogram; Nm, Newton meter; W, Watt; $r_a$ , Pearson correlation coefficient of random intercepts; $r_b$ , Pearson correlation coefficient of random slopes

## Discussion

This longitudinal observational cohort study aimed at investigating the longitudinal interaction between progressive muscle impairments and progressive gait features in growing boys with DMD. The associations between specific muscle impairments and specific gait deviations, in terms of both starting levels and progression rates, were determined. Our results provided improved insights into how specific muscle impairments contribute to specific kinematic and kinetic gait features.

The starting levels of specific muscle impairments were linked to the starting levels of specific gait deviations in boys with DMD. Our results confirm that increased anterior pelvic tilt, previously postulated as a direct effect of hip extension weakness^13,14,28^ and a compensation mechanism for knee extension weakness by positioning the ground reaction force in front of the knee joint center^1^, is indeed linked to both. Especially plantar flexion contracture, and not that much dorsiflexion weakness, was related to the drop foot in swing and subsequent decreased dorsiflexion at initial contact. This finding is consistent with Sutherland et al.^1^, who attributed the drop foot to dorsiflexion weakness early on and to plantar flexion contracture in later stages. The link between reduced hip extension moments and hip extension weakness suggests that aligning the ground reaction force more posteriorly to the hip joint center^1,15^ is indeed a compensation mechanism. Knee extension weakness was strongly related to reduced knee extension moments and knee power absorption, affirming the previously stated compensation mechanism of keeping the ground reaction force close to or in front of the knee joint center to diminish the eccentric demand on the quadriceps muscle^1,15^. Hip abduction weakness seemed to be compensated more by increased step width than by increased lateral trunk range of motion, which was suggested by Sutherland et al.^1^.The progression rates in specific muscle impairments were related to the progression rates in specific gait deviations, suggesting potential contribution. The compensation mechanism for progressive hip extension weakness by increasing the posterior trunk leaning was longitudinally confirmed^1^. This progressive hip extension weakness also resulted in progressive anterior pelvic tilt^13,14,28^. Decreasing dorsiflexion at initial contact and the progressive plantar flexion contracture interacted longitudinally. Yet, decreasing dorsiflexion at initial contact was also related to progressive knee extension weakness, confirming the previously described theory of equinus positioning as an effective compensation mechanism to align the ground reaction force in front of the knee^1^. Lastly, there was a link between increasing internal foot progression and progressive hip abduction weakness. This is rather unexpected as the ground reaction force is more medially oriented with internal foot progression and therefore this gait deviation is still not well understood^1^.

Due to the presence of multiple co-occurring impairments and their simultaneous decline over time, caution is still needed with “implying cause by association”. Detecting the causal contributors to gait pathology requires advanced computational approaches. Future studies could use machine learning with causal inference to unravel complex relationships between multiple co-occurring impairments. Additionally, physics-based modeling offers a means to directly test causality. Additionally, assessing the association between stationary and dynamic assessments might not be optimal to verify the hypothesized relationships^29^. However, establishing valid measurements of the impairments during gait is not yet possible. Furthermore, in some children with DMD, only a limited number of repeated measures were obtained. The boys with DMD enrolled in the study at varying ages, resulting in a wide baseline age range. There was also inter-subject heterogeneity due to differences in medical and clinical histories, such as clinical trial participation, adherence to night-time AFOs, periods of serial casting, functional level, etc. However, this does not influence the results, as it is expected that treatment would impact muscle impairments and gait, but not the relationship between them.

In conclusion, the starting levels and progression rates of specific muscle impairments were associated with the starting levels and progression rates of specific gait deviations in boys with DMD. This is the first study that objectively quantified these relationships, providing empirical evidence for previously proposed compensatory mechanisms and confirming several clinically assumed relationships. Moreover, novel insights were acquired that enhance our understanding of how specific muscle impairments may contribute to specific gait deviations in DMD. These findings have potential to improve clinical decision making, advance rehabilitation, orthotic and orthopedic interventions, and reveal sensitive outcome measures for assessing the efficacy of novel treatments in clinical trials.

## Data Availability

All data produced in the present study are available upon reasonable request to the authors

## List of abbreviations

3DGA: 3D gait analysis
AFOs: ankle foot orthosis
DMD: Duchenne muscular dystrophy
GC: gait cycle
MVIC: maximal voluntary isometric contraction
NMRC: Neuromuscular Reference Centre
ROM: range of motion
TD: typically developing

## Acknowledgements

The authors would like to thank all the boys and their parents who participated in this study. We also appreciate the colleagues of the University Hospital of Leuven who assisted with recruitment. A special acknowledgment goes to Elze Stoop and Tijl Dewit for the support in data collection and processing.

## Supplementary Materials

**Appendix S1:**
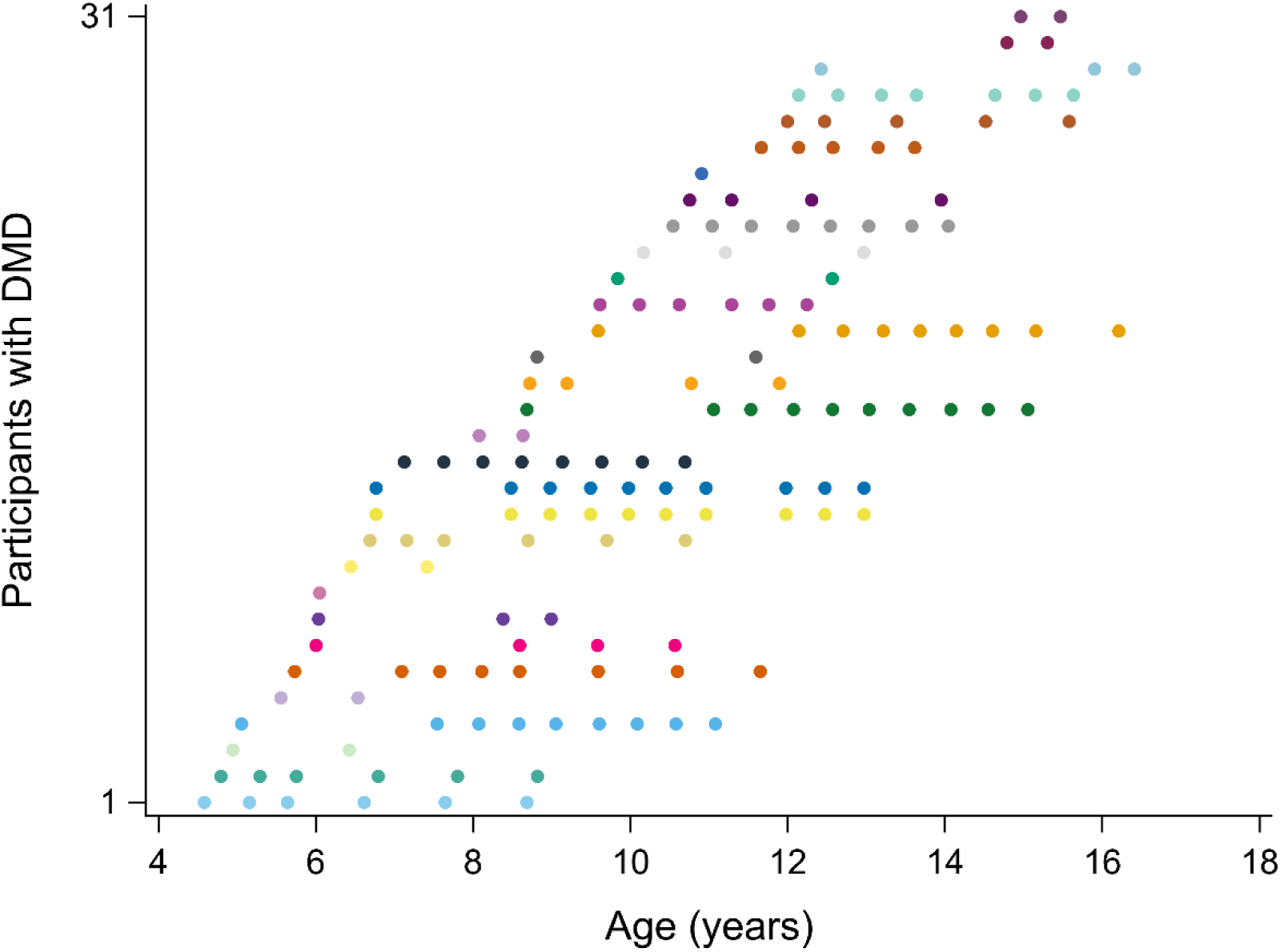
Overview of the collected longitudinal dataset. The ages at the included repeated measurements are visualized per participant with DMD, with each participant indicated by a different color. DMD, Duchenne muscular dystrophy

**Appendix S2:**
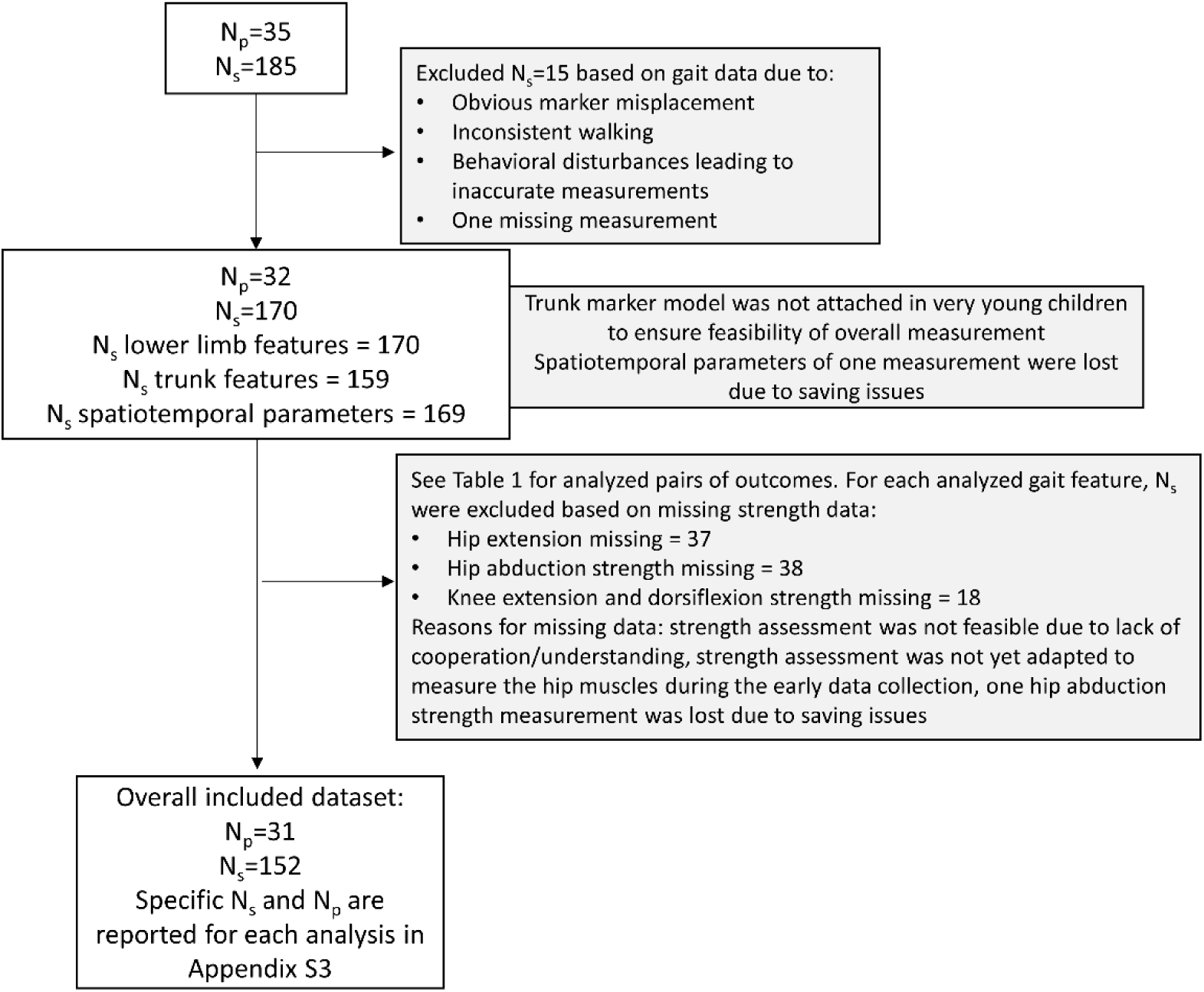
Flowchart of collected dataset, excluded data, missing data, and included dataset. N_p_, number of patients; N_s_, number of measurement sessions

**Appendix S3:** Fixed effects of joint models between gait outcomes and impairment outcomes for the boys with DMD.

| Gait | Impairments | n subj | n obs | Gait outcome |  | Impairment outcome |  |
| --- | --- | --- | --- | --- | --- | --- | --- |
| | | | | $\alpha_1$ [CI]<br>*p-value* | $\beta_1$ [CI]<br>*p-value* | $\alpha_2$ [CI]<br>*p-value* | $\beta_2$ [CI]<br>*p-value* |
| Minimal anterior trunk angle (°) | Hip extension strength deficit (z-score) | 30 | 260 | -12.3 [-15.2; -9.5]<br>**<0.0001\*** | -2.1 [-3.3; -0.9]<br>**0.0016\*** | -3.35 [-3.87; -2.84]<br>**<0.0001\*** | -0.05 [-0.32; 0.21]<br>0.6724 |
| Lateral trunk range of motion (°) | Hip abduction strength deficit (z-score) | 30 | 258 | 5.9 [4.6; 7.1]<br>**<0.0001\*** | 0.1 [-0.4; 0.7]<br>0.6234 | -2.34 [-2.78; -1.90]<br>**<0.0001\*** | -0.37 [-0.51; -0.22]<br>**<0.0001\*** |
| Maximal anterior pelvic tilt angle (°) | Hip extension strength deficit (z-score) | 31 | 266 | 18.1 [15.7; 20.5]<br>**<0.0001\*** | 1.7 [1.1; 2.3]<br>**<0.0001\*** | -3.36 [-3.86; -2.86]<br>**<0.0001\*** | -0.06 [-0.35; 0.24]<br>0.7033 |
| Maximal anterior pelvic tilt angle (°) | Knee extension strength deficit (z-score) | 31 | 266 | 18.2 [15.7; 20.6]<br>**<0.0001\*** | 1.6 [0.9; 2.4]<br>**0.0001\*** | -1.76 [-2.30; -1.22]<br>**<0.0001\*** | -0.33 [-0.50; -0.15]<br>**0.0006\*** |
| Dorsiflexion angle at initial contact (°) | Knee extension strength deficit (z-score) | 31 | 304 | -0.2 [-2.2; 1.8]<br>0.8249 | -1.4 [-2.1; -0.8]<br>**<0.0001\*** | -1.25 [-1.77; -0.73]<br>**<0.0001\*** | -0.45 [-0.56; -0.33]<br>**<0.0001\*** |
| Dorsiflexion angle at initial contact (°) | Dorsiflexion strength deficit (z-score) | 31 | 304 | -0.2 [-2.2; 1.8]<br>0.8472 | -1.4 [-2.2; -0.7]<br>**0.0003\*** | -2.31 [-2.70; -1.92]<br>**<0.0001\*** | -0.28 [-0.45; -0.12]<br>**0.0014\*** |
| Dorsiflexion angle at initial contact (°) | Dorsiflexion range of motion deficit (z-score) | 31 | 304 | -0.2 [-2.1; 1.8]<br>0.8568 | -1.4 [-2.1; -0.8]<br>**0.0001\*** | -3.03 [-3.52; -2.53]<br>**<0.0001\*** | -0.28 [-0.43; -0.12]<br>**0.0011\*** |
| Maximal dorsiflexion angle during swing (°) | Dorsiflexion strength deficit (z-score) | 31 | 304 | 2.9 [0.6; 5.3]<br>**0.0176\*** | -1.2 [-2.1; -0.3]<br>**0.0097\*** | -2.31 [-2.70; -1.92]<br>**<0.0001\*** | -0.29 [-0.46; -0.12]<br>**0.0015\*** |
| Maximal dorsiflexion angle during swing (°) | Dorsiflexion range of motion deficit (z-score) | 31 | 304 | 2.9 [0.5; 5.3]<br>**0.0203\*** | -1.2 [-1.9; -0.5]<br>**0.0017\*** | -3.00 [-3.47; -2.53]<br>**<0.0001\*** | -0.35 [-0.50; -0.20]<br>**<0.0001\*** |
| Maximal internal foot progression angle during stance (°) | Hip abduction strength deficit (z-score) | 31 | 264 | -7.7 [-10.6; -4.9]<br>**<0.0001\*** | 2.2 [0.7; 3.7]<br>**0.0057\*** | -2.35 [-2.76; -1.94]<br>**<0.0001\*** | -0.34 [-0.50; -0.18]<br>**0.0002\*** |
| Maximal hip extension moment during stance (Nm/kg) | Hip extension strength deficit (z-score) | 31 | 266 | 0.50 [0.42; 0.58]<br>**<0.0001\*** | -0.06 [-0.09; -0.03]<br>**0.0009\*** | -3.35 [-3.84; -2.85]<br>**<0.0001\*** | -0.10 [-0.36; 0.16]<br>0.4473 |
| Maximal knee extension moment during stance (Nm/kg) | Knee extension strength deficit (z-score) | 31 | 304 | 0.41 [0.36; 0.47]<br>**<0.0001\*** | -0.02 [-0.03; 0.00]<br>**0.0361\*** | -1.27 [-1.77; -0.76]<br>**<0.0001\*** | -0.41 [-0.53; -0.30]<br>**<0.0001\*** |
| Minimal knee power during burst K1 (W/kg) | Knee extension strength deficit (z-score) | 31 | 304 | -0.70 [-0.89; -0.51]<br>**<0.0001\*** | 0.05 [0.01; 0.09]<br>**0.0200\*** | -1.26 [-1.77; -0.75]<br>**<0.0001\*** | -0.41 [-0.53; -0.30]<br>**<0.0001\*** |
| Normalized step width (/) | Hip abduction strength deficit (z-score) | 30 | 262 | 0.28 [0.26; 0.29]<br>**<0.0001\*** | 0.01 [0.00; 0.03]<br>**0.0248\*** | -2.33 [-2.74; -1.91]<br>**<0.0001\*** | -0.35 [-0.51; -0.18]<br>**0.0003\*** |

Each row is one joint model.
*p-values in bold indicate significance level at $p < 0.05$ .*
The following symbols represent: $\alpha_1$ , intercept of gait outcome; $\beta_1$ , regression slope of time for gait outcome; $\alpha_2$ , intercept of impairment outcome; $\beta_2$ , regression slope of time for impairment outcome.
CI, 95% confidence interval; DMD, Duchenne muscular dystrophy; kg, kilogram; n, number; Nm, Newton meter; o, observations; subj, subjects
W, Watt;

**Appendix S4:** Covariance matrix of random effects and measurement error of joint models between gait outcomes and impairment outcomes for the boys with DMD.

| Gait | Impairments | Covariance matrix |  |  |  |  |  |  |  |  |  | Measurement error |  |
| --- | --- | --- | --- | --- | --- | --- | --- | --- | --- | --- | --- | --- | --- |
| | | $d_{a1a1}$ | $d_{a1a2}$ | $d_{a2a2}$ | $d_{a1b1}$ | $d_{a2b1}$ | $d_{b1b1}$ | $d_{a1b2}$ | $d_{a2b2}$ | $d_{b1b2}$ | $d_{b2b2}$ | $\varepsilon_1$ | $\varepsilon_2$ |
| Minimal anterior trunk angle (°) | Hip extension strength deficit (z-score) | 54.971 | 3.477 | 1.551 | 6.609 | 2.149 | 6.468 | 0.938 | 0.074 | 0.653 | 0.192 | 4.719 | 0.578 |
| Lateral trunk range of motion (°) | Hip abduction strength deficit (z-score) | 8.427 | -0.575 | 1.129 | -0.589 | -0.244 | 0.688 | -0.584 | 0.064 | 0.118 | 0.072 | 3.065 | 0.599 |
| Maximal anterior pelvic tilt angle (°) | Hip extension strength deficit (z-score) | 40.164 | -4.124 | 1.434 | 4.192 | -0.400 | 1.641 | -0.267 | 0.068 | -0.431 | 0.171 | 6.522 | 0.582 |
| Maximal anterior pelvic tilt angle (°) | Knee extension strength deficit (z-score) | 40.196 | -4.213 | 1.998 | 3.509 | -0.445 | 1.478 | 0.007 | 0.003 | -0.195 | 0.094 | 6.577 | 0.220 |
| Dorsiflexion angle at initial contact (°) | Knee extension strength deficit (z-score) | 22.289 | 1.253 | 1.756 | 1.203 | 0.259 | 0.855 | 0.606 | 0.047 | 0.154 | 0.059 | 11.197 | 0.364 |
| Dorsiflexion angle at initial contact (°) | Dorsiflexion strength deficit (z-score) | 22.322 | 1.998 | 0.732 | 0.445 | -0.019 | 0.966 | 0.588 | 0.013 | 0.156 | 0.079 | 11.056 | 0.608 |
| Dorsiflexion angle at initial contact (°) | Dorsiflexion range of motion deficit (z-score) | 22.037 | 4.884 | 2.023 | -0.672 | 0.000 | 1.424 | 0.000 | -0.131 | 0.182 | 0.059 | 10.885 | 0.566 |
| Maximal dorsiflexion angle during swing (°) | Dorsiflexion strength deficit (z-score) | 35.810 | 2.516 | 0.725 | -0.776 | 0.139 | 1.473 | 0.650 | 0.020 | 0.130 | 0.077 | 10.008 | 0.605 |
| Maximal dorsiflexion angle during swing (°) | Dorsiflexion range of motion deficit (z-score) | 35.740 | 4.445 | 1.258 | -0.543 | 0.294 | 1.453 | 1.086 | 0.079 | 0.122 | 0.061 | 9.952 | 0.578 |
| Maximal internal foot progression angle during stance (°) | Hip abduction strength deficit (z-score) | 52.003 | -2.017 | 0.922 | 3.493 | -0.817 | 7.233 | 0.755 | 0.062 | -0.435 | 0.087 | 11.410 | 0.623 |
| Maximal hip extension moment during stance (Nm/kg) | Hip extension strength deficit (z-score) | 0.028 | 0.108 | 1.470 | 0.059 | -0.106 | 0.264 | / | / | / | / | 0.027 | 0.582 |
| Maximal knee extension moment during stance (Nm/kg) | Knee extension strength deficit (z-score) | 0.024 | 0.142 | 1.693 | 0.008 | 0.050 | 0.048 | / | / | / | / | 0.015 | 0.372 |
| Minimal knee power during burst K1 (W/kg) | Knee extension strength deficit (z-score) | 0.275 | -0.519 | 1.708 | -0.039 | 0.045 | 0.050 | / | / | / | / | 0.107 | 0.370 |
| Normalized step width (/) | Hip abduction strength deficit (z-score) | 0.001 | -0.019 | 0.952 | 0.000 | -0.008 | 0.000 | -0.003 | 0.084 | -0.002 | 0.093 | 0.001 | 0.616 |

Each row is one joint model.
The following symbols represent: $a_1$ , random intercept for gait outcome; $b_1$ , random slope of time for gait outcome; $a_2$ , random intercept for impairment outcome; $b_2$ , random slope of time for impairment outcome; $d_{a_1a_1}$ , variance random intercept $a_1$ ; $d_{a_1a_2}$ , covariance random intercepts $a_1$ and $a_2$ ; $d_{a_2a_2}$ , variance random intercept $a_2$ ; $d_{a_1b_1}$ , covariance random intercept $a_1$ and random slope $b_1$ ; $d_{a_2b_1}$ , covariance random intercept $a_2$ and random slope $b_1$ ; $d_{b_1b_1}$ , variance random slope $b_1$ ; $d_{a_1b_2}$ , covariance random intercept $a_1$ and random slope $b_2$ ; $d_{a_2b_2}$ , covariance random intercept $a_2$ and random slope $b_2$ ; $d_{b_1b_2}$ , covariance random slopes $b_1$ and $b_2$ ; $d_{b_2b_2}$ , variance random slope $b_2$ ; $\varepsilon_{1ij}$ , measurement error of gait outcome; $\varepsilon_{2ij}$ , measurement error of muscle impairment outcome;
DMD, Duchenne muscular dystrophy; kg, kilogram; Nm, Newton meter; W, Watt;

